# Epidemic Situation and Forecasting if COVID-19 in Saudi Arabia using SIR model

**DOI:** 10.1101/2020.05.05.20091520

**Authors:** Haitham Khoj, Alaa F Mujallad

## Abstract

**Background:** Saudi Arabia is one of the countries affected by COVID-19 pandemic. This will lead to negative impacts in many sectors. Saudi Arabia not only plays an important role on the economical side because it is the leading country in oil production, but also because it is considered the heart of the Islamic countries. Although protective measures have been implemented in Saudi Arabia, the number of COVID-19 cases has increased.

**Aims of the study:** This study aimed to employ SIR model to forecast the peak of COVID-19 progression and an estimation of it is end in Saudi Arabia.

**Method:** Based on the World Health Organization data on COVID-19 progression in Saudi Arabia from March 3rd to April 29th, 2020, we reliably estimate the constant parameters and make predictions on the inflection point and potential ending time. Susceptible, Infected, and Recovered are the main components of the SIR model that were used to run the analysis.

**Result:** The data showed an interesting result about the peak of the disease progression. It is projected to occur around the 20th day after running the model. According to the model, the peak time will be around the 20th of May. Then the cases will decrease until the 55th day, which is around June 20th.

**Conclusion:** The result predicts a second peak and an estimation end of COVID-19 in Saudi Arabia. This data can inform the policy makers, who should try to contain the virus, to be prepared for what is coming next.

Key Messages:

## 1. Implications for policy makers

Healthcare providers and economists engage in politics and policy-making to achieve public health outcomes. Findings from this proposed study will inform policy-makers as they strive to raise awareness about COVID-19. The findings of this study will show the policy-makers to what degree COVID-19 will affect the health of people in Saudi Arabia. The policy makers will be motivated to implement policies and procedures for national healthcare that will allow Saudi officials to be prepare for this pandemic.

## 2. Implications for public

There is a lack of evidence regarding estimation time of COVID-19 progression, among Saudi Arabian population. By addressing this knowledge gap, this study will provide evidence with significance for the public to adhere to COVID-19 protective measures that will have a positive impact to slow down the spread of the disease.

### Background

With the emergence of COVID-19 and the World Health Organization (WHO) declaring COVID-19 a global pandemic, many have been left questioning what COVID-19 is and how long it is going to last. When will we be able to contain COVID-19? When will the peak and the end to this pandemic occur? Answers to these questions are essential for implementing the policies needed, especially in the economic and healthcare sectors. The whole world is affected, and many have lost their lives; hospitals are packed, healthcare providers are overwhelmed, and businesses are closed.

The pathogen of SARS-CoV-2 has the same phylogenetic similarity to SARS-CoV and MERS-CoV. Coronavirus can infect humans and some wild animals such as bats, birds, and mice. These viruses affect the major systems in the body which are respiratory, gastrointestinal, hepatic, and central nervous system. The outbreak of coronaviruses demonstrated evidence of animal-to-human transmission and human-to-human transmission. The vaccines and appropriate treatment are needed to stop the COVID-19 pandemic and any upcoming coronavirus outbreaks in the future. As long as there is direct contact between humans and animals, and the changes of the climate and ecology, there is still a possibility that another pandemic may occur (Chen, 2020). Saudi Arabia is one of many countries suffering from COVID-19. This will lead to negative impacts in many sectors. Saudi Arabia is a premier economic power, not only because it is the leading country in oil production, but also because it is considered the heart of the Islamic countries. It is the country that holds Hajj, the largest annual gathering in the world, and a very important event for every Muslim. Saudi Arabia receives 3 million pilgrims yearly who perform Hajj, and 7.5 million people to do Umrah from more than 180 countries. Umrah is a smaller ritual comparing to Hajj, performed year-round (Yezli, et al., 2016). China is one of the countries who is included in the visa program to visit Saudi Arabia to preform religious rituals. What we have to be concerned about is that around 46% of the pilgrims are over 56 years of age and 50% of them have pre-existing-medical conditions (Shahul & Memish, 2020.) Making advanced preparations to receive this many people is essential, and it will be even more necessary in order to make sure COVID-19 precautions are followed (Ascoura, 2013).

One of the characteristics of COVID-19 is that it is highly contagious; this means it can spread very fast. The total confirmed cases worldwide is 3,267,184 with 229,971 deaths (WHO) as of May 2, 2020. Although protective measures have been implemented in Saudi Arabia, such as countries locking down, people practicing social distancing, and applying the infection control precautions (MOH), the number of cases has increased. The total confirmed cases worldwide is 24,097 with 169 deaths (WHO) as of May 2, 2020 (WHO).

The Saudi government applied early measures that included suspending this year’s Umrah pilgrimage on the 27^th^ of February (Shahul el al, 2020), locking down most of the affected cities in Saudi Arabia. However, the number of COVID-19 cases is increasing. Data from China can be used to project the spread of COVID-19 in Saudi Arabia, which managed to have less cases of COVID-19 in March, a mathematical calculation could provide overview or estimation about the spread of COVID-19 in Saudi Arabia.

SARS-CoV-2 is not the only infectious agent that made negative impacts on humans. Through the centuries, numerous agents attacked humans and led to critical cases and deaths. Europe had an attack from the Black Death in the fourteen century, which lead to deaths for a third of the populations. Moreover, in the USA, Yellow Fever has taken lives of 5000 people out of 50000. Models for infectious disease have been used to forecast the spread of diseases, and they were successful. SIR model is one of the most common models that could forecast the end point of COVID-19 in Saudi Arabia (Bailey, 1975).

Susceptible, Infected, and Recovered (SIR) is a commonly used model that tracks the process of a certain disease. SIR is used to estimate vital epidemiological parameters, such as forecasting works and transmission rates (Ming, 2020). Since some people are asymptomatic, it is difficult to trace this virus and to determine the first case in Saudi Arabia. Moreover, the uncertainty of determining the exact number of infected people is due to some of them being infected but either asymptomatic or have mild symptoms, such as a common cold, will be a problem too. It is interesting to use a mathematical procedure that has dealt with diseases, such as COVID-19, and apply it to have an evidence basis for forecasting.

## Method

Based on the nature of COVID-19 and the characteristics of the virus, SARS-CoV-2 that causes the disease, SIR model is a good fit to forecast the end period of COVID-19. A mathematical model is an important component in our research because it is the foundation that can guide scholars to identify which concepts are important, what these concepts are like, and how they may be related to each other. They shape the plan for the future, and then influence how the findings are interpreted. Susceptible, infected, and recovered model (SIR) is a very common model that is used in forecasting the disease progression of many infectious diseases.

The model first was presented by Kermack and Mckendrick (1927), in our study we employ the SIR model to predict the peak of the COVID-19 progression and provide a rough estimation of its end. The growth index will be calculated, according to the data from March 2 until April 29. Susceptible, Infected, and Recovered are the main components of the model. The susceptible will be the whole population of the country of Saudi Arabia, which is roughly 34,000,000, because this disease is highly contagious, this data was taken from an authoritative source. The infected and the recovered are secondary data that have been taken from Worldometer.

So, a population of N in our study divided to (S) susceptible, (I) infected, and (R) recovery
where

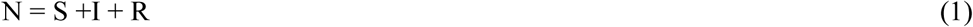

So, we can rewrite it as a function of time as follows:

S = S(t) number of susceptible individuals over time, I = I(t) number of infected individuals over time, and R = R(t) number of recovered individuals over time.

Thus, Susceptible differential equation

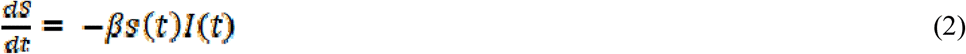

The Recovered differential Equation. r(t)

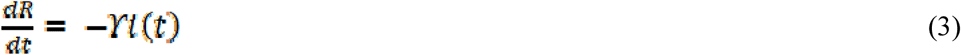

At the initial condition at initial time {S(0), I(0), R(0)} where parameter β is the infection rate and parameter β the recovery rate.

Parameter β represent the rate of Susceptible who get infected over time, and parameter β represent the recovery rate whose been infected.

By the running the simple SIR model we can forecast when COVID-19 will slowdown in Saudi Arabia over time where t = 70.

## Result

This section will present the results for our SIR calculations. Our time path was 70 days as of the study in China calculated their based on 70 days (Pan, et al., 2020). We assumed at t = 0 we have S(0) = .99 percent of the population is Susceptible and starting infected at I(0) = 0.01, which was one percent of population infected, and recovery rate is zero R(0) = 0. We compared our results with China cases, and it showed similar results to the constant parameters for SIR model that have been used to forecast the epidemic COVID-19 in China. Table 1 showed the constant parameters for SIR model in Saudi Arabia. One important policy Saudi Arabia has implemented is lockdown of all activities and stay at home order which we did not measure to our study.

**Table 1:**
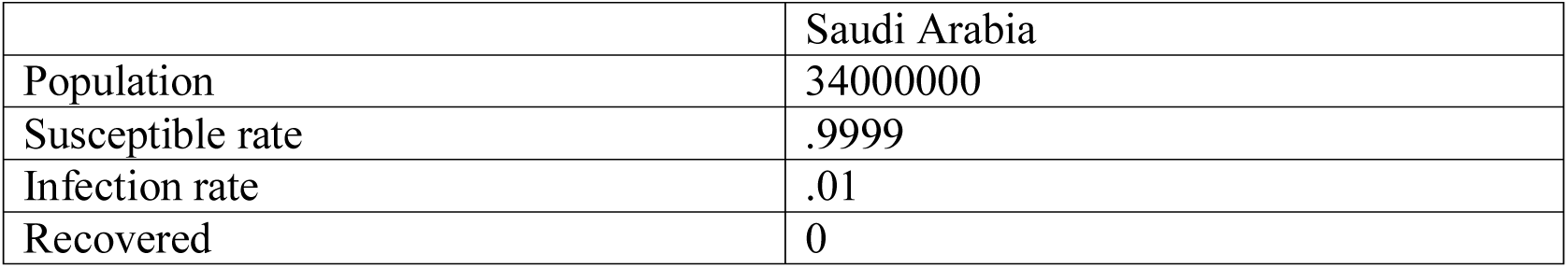
Summary of constant parameters for SIR model to forecast the epidemic COVID-19 in Saudi Arabia

|  | Saudi Arabia |
| --- | --- |
| Population | 34000000 |
| Susceptible rate | .9999 |
| Infection rate | .01 |
| Recovered | 0 |

The data showed the daily number of infected people increase with COVID-19 starting from 3/2/2020 to 4/27/2020, and the number of recovery patients with a recovery rate of 13%. Starting from mid-April the daily number of infected increased dramatically, while recovery rates are still low compared the large increased in the infected number as shown in graph 1. Economically it indicates that the cost and pursuer that the economy can face as number of people infected and recovery rate is low too high.

**Graph 1:**
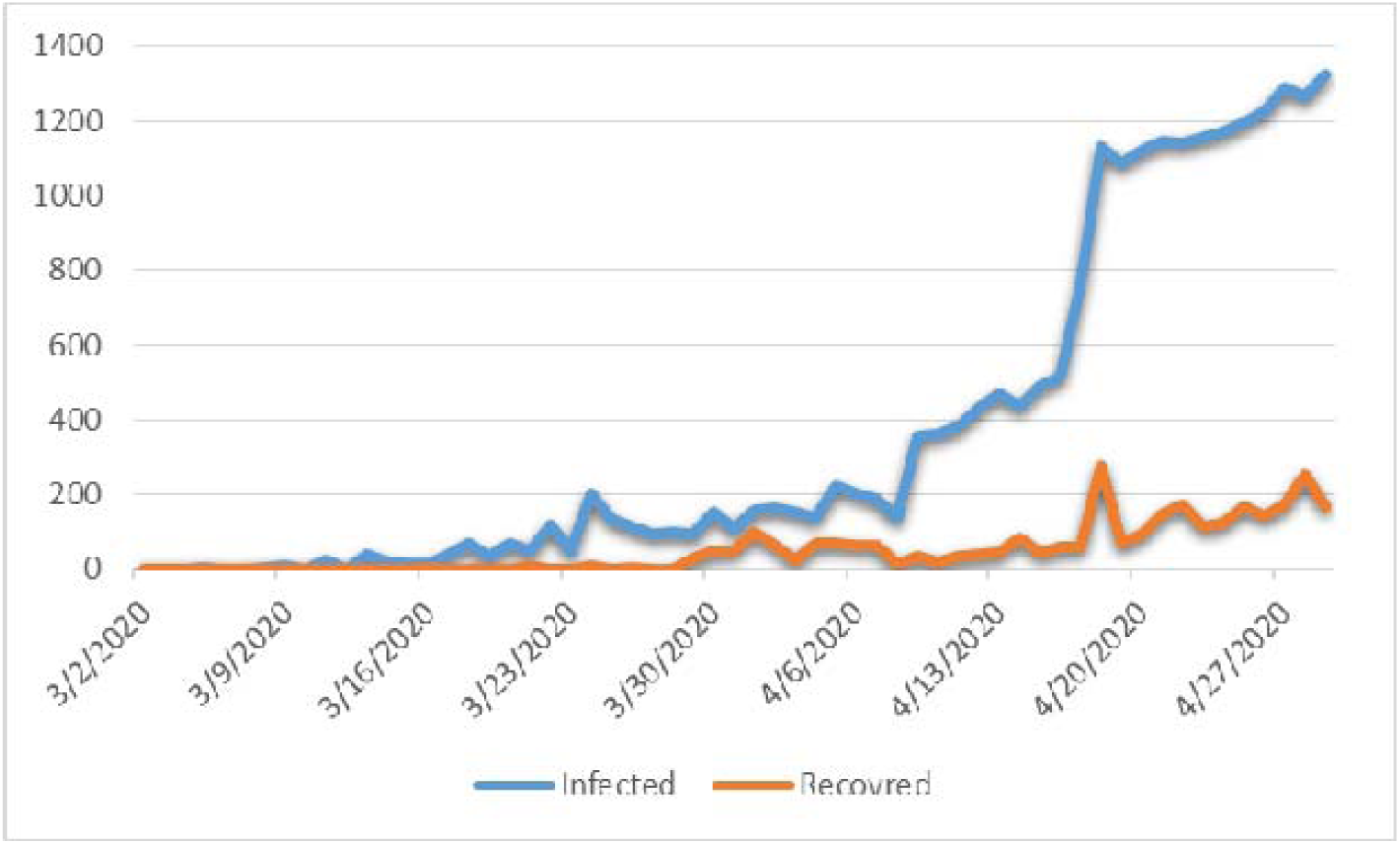
Number of infected VS number of recovered daily

According to the growth index of COVID-19 over time, where it’s clear that the increase in number of cases. Timeline clearly indicates a growth in the case index since the first case and its rising but in sustainable rate, which is shown in graph 2.

**Graph 2:**
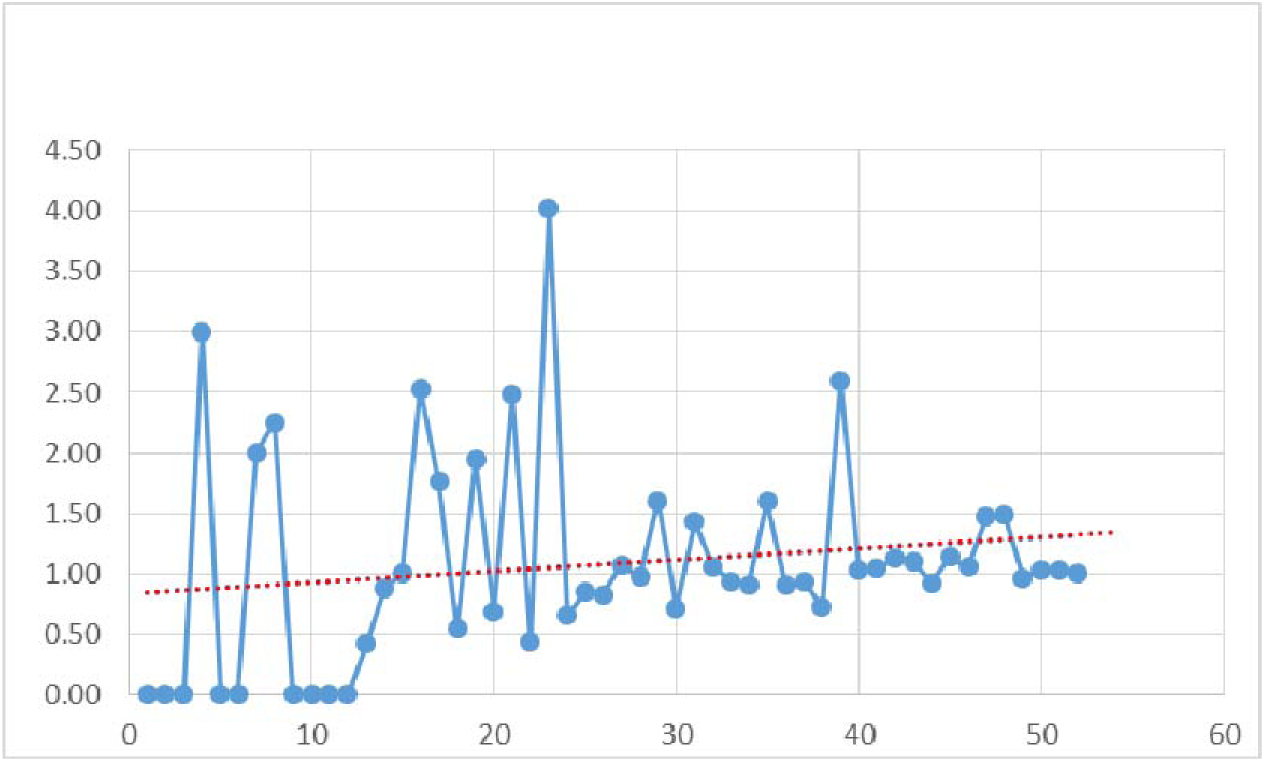
Growth Index of COVID-19

The data showed an interesting result about the peak of the disease progression. It is projected to occur around the 20^th^ day after running the model. According to the model, the peak time will be around the 20^th^ of May. Then the cases will decrease until the 55^th^ day, which is around June 20^th^.

**Graph 3:**
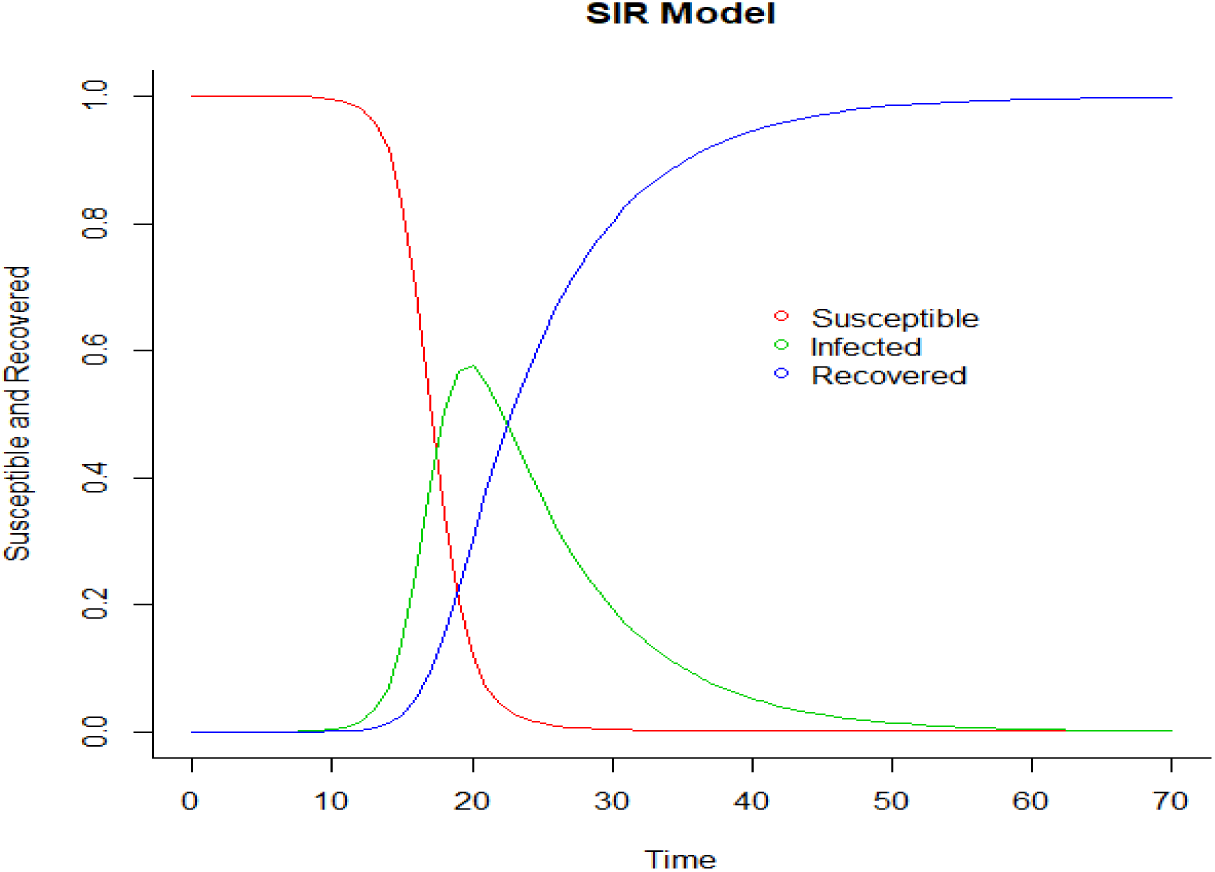
Modeling the Spread of the Disease

According to graph 2, we have data for 55 number of days since the emergence of the first infected person in Saudi Arabia which was on March 3^rd^. The parameters have been used according to those data. The model and the parameters allowed us to forecast the time needed to predict the end time of COVID-19. The other countries that had declines took around three months. So, the time was estimated around 70 days from the start date of 29 April.

## Discussion

COVID-19 left us uncertain about the nature of the disease, and what will happen in the future. The uncertainty of the disease not only impacts the health sector, but the economy as well. By analyzing the trend of this virus across other countries, the SIR model gave us an optimistic picture about an estimated time to end this pandemic. To make the number of cases less than the hypothetical result, people in Saudi Arabia must follow the same protective measures to prevent the spread of the virus. Health officials implemented productive measures such stay at home orders and ministry of finance have a stimulus package around SAR 120 billion to lessen the Impact of COVID-19 on Economic Activities and Private Sector (Ministry of Finance). This study helps policy makers in considering further action as our model predicts the slowness of spread of COVID-19 and the volume of any stimulus package needed.

In the study, SIR model using R software package was employed to predict an end point of COVID-19 in Saudi Arabia. Previous studies have investigated an outbreak of infectious disease and its severity using SIR in different countries and exploring different approaches. For example, COVID-19 outside Hubei in China was less intense in another other region compared to Hubei (Lili Wang et al. 2020).

Based on the data above, the peak time will affect around 40% of the Saudi population. As a result of this trend, the peak time will be the most dangerous time for COVID-19 transmission. Prolonged social distancing is needed to prevent catastrophic events in the healthcare system. According to this model simulation, urgent actions are needed to prepare for any critical situations in the healthcare and economic sectors.

People must take precaution seriously to end the COVID-19 pandemic and heed the statistics available. The government did great to help citizens and non-citizens there by providing them with the care that they need and overcomes their financial losses. In addition, healthcare providers were privileged by postponing all the debts as an appreciation for their hard work. The Saudi Arabia health officials have already implemented strong predictive measures to reduce the spread of the virus such as stay at home order, closing malls, etc. The infection rate in Saudi Arabia as of today is .93 which means every infected person will infect roughly one. This is still a good number to make it easier for health care providers to combat the COVID-19. Therefore, there will be success in delaying and eliminating a sudden increase of cases of COVID-19. Finally, the study suggests a second wave of the COVID-19 will start in 20 to 30 days before it slows down again in Saudi Arabia.

## Conclusion

SIR model has a proven history for forecasting the spread of many infectious diseases. After the application of SIR model to forecast COVID-19 In Saudi Arabia, the result predicts a second peak and an estimation end of COVID-19 in Saudi Arabia. Data can inform the policy makers, who should try to contain the virus, to be prepared for what is coming next. Also, people must continue their home quarantine and social distancing to reduce circumstances of COVID-19 whether economically or in the healthcare sector. Recently, Saudi Arabia health officials took preventive and defensive measures to slowdown the spread, and buy time until health scientists find a vaccine or cure of the disease. Further research will be investigating the effectiveness of the measure implemented by health officials in Saudi Arabia.

## Data Availability

World Health Organization

